# Shoulder injuries in elite female cricket players: Insights from 8 seasons

**DOI:** 10.1101/2025.01.15.25320632

**Authors:** Gordon Pritchard, Pallavi Deshmukh, Anna E. Saw, Kate Beerworth, Kevin Sims

## Abstract

**Objectives:** To describe the incidence, prevalence, characteristics, and management of shoulder injuries requiring medical attention in elite female cricket players.

**Design:** Retrospective cohort.

**Setting:** Australian state, territory, and national cricket teams between July 2015 and June 2023.

**Participants:** Elite female cricket players.

**Independent variables:** Medical attention and general time-loss shoulder injuries.

**Main outcome measures:** Incidence, prevalence, characteristics, recovery time-frames, activity modifications.

**Results:** 409 shoulder injuries were recorded, with an average incidence of 12.9 per 100 players per season. Gradual onset injuries were most commonly experienced by pace bowlers (51%). Sudden onset injuries were most commonly sustained whilst fielding (69%). Four in five injuries did not require the player to be unavailable to play or train. Modified activities (e.g., throwing, diving, bowling) were typically required for between 2 weeks and 6 months. Players typically returned to full unrestricted match play between 1-8.5 months, longer for recurrent injuries (p=0.007).

**Conclusions:** Shoulder injuries are a considerable burden in elite female cricket players. Despite only one in five injuries resulting in a player being unavailable to play or train, impaired shoulder function may reduce the overall performance of the player and the team. Risk reduction strategies may be targeted to at risk players (pace bowlers) and activities (throwing and diving) to reduce the burden of shoulder injuries in this cohort. Further consideration may also be given to management strategies to reduce the risk of exacerbations and recurrent injuries which may prolong recovery.

## Introduction

Shoulder injuries are common in athletes who throw overhead, and dive and land with an outstretched arm.^1^ Cricket involves diving to field the ball and throwing to return the ball to the centre wicket. Throws include shorter fast precision-based throws at the stumps and long throws from the outfield. Cricket players who bowl do so at a much higher volume than throwing, with the bowling action involving shoulder abduction and rotation.

To quantify the burden of shoulder injuries in cricket, the definition of injury must be clearly understood. The International consensus statement on injury surveillance in cricket (2016) defines a match time-loss injury as an injury which results in a player missing a match they would otherwise have participated in; whereas a general time-loss injury results in a player being deemed unfit to play a match, regardless of whether an actual match was missed.^2^ Medical attention injuries include time-loss and non time-loss injuries which require medical attention and may affect the player’s ability to train or play, but do not necessarily prevent the player from participating in training or matches.^2^

Shoulder injuries resulting in match time-loss are reported to occur at an average annual incidence of 3.1 injuries per 100 elite male cricket players in England and Wales.^3^ A similar rate has been reported in elite male and female cricket players in Australia (3.8 and 2.8 shoulder and upper arm injuries per 100 male and female players respectively).^4^ In the same elite Australian cohort, the annual incidence of medical attention injuries was 25.3 and 36.9 per 100 male and female players respectively (shoulder and upper arm).^4^ Female international development players in England and Wales reported no match time-loss shoulder injuries over four years, but an average of 12 medical attention injuries per 100 players.^5^

Whilst there is heterogeneity in cohorts and methods in these and other studies reporting shoulder injuries in elite cricket players,^6–9^ there are two key themes: firstly, the rate of medical attention shoulder injuries is high in elite cricket players, considerably higher than time-loss injuries, and; secondly, the rate of medical attention shoulder injuries in elite female players may be higher than elite male players, although there is less data in this cohort. To understand the burden of medical attention shoulder injuries, research is needed to quantify the type and duration of activity modifications required to enable a player to continue to participate in training and matches.

In addition to improving understanding of the burden of shoulder injuries in elite female cricket players, there is a need to understand which players are most affected by shoulder injuries, and which scenarios pose a higher risk. This information may be used to guide risk reduction strategies. Previous research has inconsistently described which cricket players are most at risk (batters^9^, fast bowlers^10,11^) and which activities are a higher risk for shoulder injury (fielding^7,10,12–14^, fast bowling^15^). In addition to the heterogeneity of studies mentioned previously, a key factor to these inconsistent findings may be the grouping of gradual onset and sudden onset injuries.

Therefore, the purpose of this study is to describe the incidence, prevalence, and nature of shoulder injuries in elite female cricket players, with distinction for medical attention and general time-loss injuries, and gradual onset and sudden onset injuries. The second purpose is to describe the management of shoulder injuries in elite female cricket players, including modifications to cricket-specific skills and activities; and time to return to key activities.

## Methods

Ethics approval was attained from the La Trobe University Human Research Ethics Committee (HEC20058). Individual consent was waived as data was collected as part of routine athlete servicing and is not identifiable in this research. Data were retrieved from Cricket Australia’s online database (Athlete Management System, Fair Play Pty Ltd.).

1019 Australian female cricket players participated in elite senior and emerging state and national cricket programs over 8 seasons (1 July 2015-30 June 2023). The average number of players each season was 396, and the average age of players at the start of each season was 19.0 ± 5.5 years.

All medical attention injuries were included if they were coded with a) a diagnosis related to the shoulder complex, and b) occurred during cricket participation. For the purpose of this study, the shoulder complex was defined as intra- and extra-articular structures on the glenohumeral and acromio-clavicular joints, along with the rotator cuff and long head of biceps tendon. Injuries were excluded if the injury was beyond the defined shoulder complex (e.g., latissimus dorsi, triceps, and biceps muscles) or the injury was not related to cricket training or match participation.

Injuries were diagnosed by team medical staff (doctor, physiotherapist) from clinical assessment, supported by radiological assessment if clinically indicated. Injuries were coded by the treating practitioner using the Orchard Sports Injury Classification System.^16^ Diagnoses were grouped as the following umbrella diagnoses for analysis: Rotator cuff related shoulder pain (internal impingement, subacromial impingement, impingement/synovitis, rotator cuff muscle injury, subscapularis tendon injury, supraspinatus tendon injury, infraspinatus tendon injury), shoulder instability and glenohumeral joint (shoulder dislocation, shoulder subluxation, instability, glenohumeral ligament, glenohumeral joint chondral injury, or labral lesion), other non-rotator cuff shoulder muscle injury (deltoid, pectoralis major, contusion), acromioclavicular joint (acromioclavicular joint sprain or contusion or synovitis), biceps tendon pathology (biceps tendon), acute shoulder fracture (clavicle), and undiagnosed shoulder injury (shoulder injuries, shoulder pain undiagnosed).

A medical attention injury was considered a repeat injury if the player had an injury to the same side and with the same diagnosis group recorded previously during the study period, which had been closed off (unrestricted return to play) and a new injury entry made for the repeat injury. A general time-loss injury was classified as recurrent if the player had the same time-loss injury to the same side recorded previously and had successfully returned to match availability, or the medical staff had coded it as a recurrent injury from the player’s history (previous injury preceded the study period). An exacerbation was classified if the player had not returned to match availability before regressing in the degree of modification required.

Clinical consultation notes were reviewed to determine participation status and details of any modifications to usual training and/or match participation. Dates ceasing and commencing training and playing were verified by cross-referencing with activity data entered by staff and players. Participation status were classified as:

A. Full participation: Player able to fully participate in training and available for match selection if a match was scheduled that day.
B. Modified participation: Player restricted to modified activities on one or more cricket-related activities (match, skills training, strength and conditioning). Available for match selection (with modified activity) if a match was scheduled that day (non-time-loss).
C. No participation: Unable to train and unavailable for match selection if a match was scheduled that day (time-loss).

Descriptive statistics were used to characterise the players and injuries included in the cohort. Details pertaining to less than 5 cases were not explicitly reported, as per ethics requirements. Proportions were calculated with a Wilson 95% confidence interval (CI). Injury incidence was calculated as the average number of injuries per 100 players per season. Injury prevalence was calculated as the percentage of players unavailable or restricted to modified activities on any given day. Independent samples median tests were used to compare measures of burden between injuries with different onset (gradual, sudden), diagnoses of predominant injuries (rotator cuff related shoulder pain, shoulder instability and glenohumeral joint), and recurrence. Analyses were conducted in Microsoft Excel (Microsoft 365, Microsoft Corporation, Washington, USA) and SPSS (version 28 IBM, Armonk, NY, USA).

Qualitative content analysis was used to understand the treatment and management of shoulder injuries, including short-term and ongoing modifications to activities. Content analysis was completed by one author who was independent of the clinical management of players (AS). Themes were then reviewed and clarified by the research team familiar with the clinical management of players and extracted data (GP, PD, KB).

## Results

409 injuries from 204 individuals met the inclusion criteria (42 players had 2 injuries, 17 players had 3 injuries, 30 players had 4 or more injuries). General time-loss injuries accounted for 80 injuries. The average age of players at the time of their first medical attention injury was 20.8 ± 5.0 (range 13-35) years.

Approximately half of all medical attention injuries were sustained by players whose dominant skill included pace bowling (pace bowlers and all rounder pace bowlers) (50%, 95% confidence interval 45-54%). The other half of injuries was largely contributed by batters, spin bowlers, and all rounder spin bowlers.

The average incidence of medical attention shoulder injuries was 12.9 per 100 players per season. The average incidence of general time-loss injuries was 2.5 per 100 players per season. The average prevalence (the number of players being unavailable to play in a match at any given time of the season) was 0.4%, and the average prevalence of players restricted to modified cricket-related activities (but able to play) was 2.3% per season.

Gradual onset injuries occurred throughout the season, with a larger proportion of medical attention injuries mid-season (40%, 33-36%) and a larger proportion of general time-loss injuries pre-season (41%, 22-64%). Gradual onset injuries were more commonly experienced by a pace bowler, to their dominant side, and with a diagnosis of rotator cuff related shoulder pain. Full details and statistics for gradual onset shoulder injuries are presented in Table 1.

**Table 1.** Characteristics and burden of gradual onset shoulder injuries sustained by elite Australian female cricket players over 8 seasons.

|  | **Medical attention (n=220)** |  |  | **General time-loss (n=17)** |  |  |
| --- | --- | --- | --- | --- | --- | --- |
|  | **Proportion of cases (%, 95% CI)** | **Average incidence / 100 players / season** | **Prevalence modified participation (% players restricted on any given day)** | **Proportion of cases (%, 95% CI)** | **Average incidence / 100 players / season** | **Prevalence no participation (% players unable to play on any given day)** |
| **Time of season<sup>1</sup>** |  |  |  |  |  |  |
| Pre-season (May-July) | 20 (15-26) | 1.4 | 9.4 | 41 (22-64) | 0.2 | 0.9 |
| Early-season (August-October) | 26 (21-32) | 1.8 | 12.5 | 6 (1-27) | 0.0 | 0.0 |
| Mid-season (November-January) | 40 (33-46) | 2.7 | 15.8 | 29 (13-53) | 0.2 | 0.8 |
| Late-season (February-April) | 15 (11-20) | 1.0 | 5.7 | 24 (10-47) | 0.1 | 1.0 |
| **Dominant skill of player** |  |  |  |  |  |  |
| Batting | 26 (21-32) | 1.8 | 4.2 | 24 (10-47) | 0.1 | 0.2 |
| Pace bowling or Pace bowling all rounder | 51 (45-58) | 0.3 | 4.3 | 47 (26-69) | 0.3 | 0.2 |
| Spin bowling or Spin bowling all rounder | 19 (14-24) | 0.2 | 2.1 | 29 (13-53) | 0.2 | 0.3 |
| Wicket keeping or wicket keeping all rounder | 4 (2-8) | 0.0 | 0.2 | 0 (0-18) | 0.0 | 0.0 |
| **Injury side** |  |  |  |  |  |  |
| Dominant | 82 (77-87) | 5.7 | 9.6 | 88 (66-97) | 0.5 | 0.4 |
| Non dominant | 18 (13-23) | 1.2 | 1.3 | 12 (3-34) | 0.1 | 0.2 |
| **Diagnosis** |  |  |  |  |  |  |
| Rotator cuff related shoulder pain | 39 (32-45) | 2.7 | 5.3 | 59 (36-78) | 0.3 | 0.4 |
| Shoulder instability and glenohumeral joint | 22 (17-28) | 1.5 | 4.8 | 41 (22-64) | 0.2 | 0.3 |
| Other shoulder muscle injury (non rotator cuff) | 29 (23-35) | 2.0 | 0.5 | 0 (0-18) | 0.0 | 0.0 |
| Acromioclavicular joint | 3 (1-6) | 0.2 | 0.1 | 0 (0-18) | 0.0 | 0.0 |
| Biceps tendon pathology | 5 (3-9) | 0.4 | 0.2 | 0 (0-18) | 0.0 | 0.0 |
| Undiagnosed shoulder injury | 2 (1-5) | 0.2 | 0.0 | 0 (0-18) | 0.0 | 0.0 |

1 Time of season reflects Australian domestic season, however may not apply to players participating in international competitions.

Sudden onset injuries occurred throughout the season, with a larger proportion of both medical attention and general time-loss injuries mid-season (39% (32-46%) and 40% (29-52%) respectively). Injuries occurred during matches (48%, 41-55%), cricket training (42%, 35-49%), strength training (8%, 5-13%), and warm-up (2%, 1-5%). Sudden onset injuries more commonly occurred whilst fielding, to both dominant and non-dominant sides. The primary mechanism of injury whilst fielding or wicket-keeping was catching/diving/falling (60%, 51-67% of fielding injuries) followed by throwing (35%, 27-43%). Ball collision (impact) resulted in 4% (2-8%) sudden onset injuries, affecting batters and fielders (including bowlers). Rotator cuff related shoulder pain contributed the largest proportion of medical attention injuries (40%, 33-47%), whilst shoulder instability and glenohumeral joint injuries contributed the largest proportion of general time-loss injures (44%, 33-57%). Full details and statistics for sudden onset shoulder injuries are presented in Table 1.

Repeat injuries accounted for 19% (16-23%) of all medical attention injuries, occurring to the same player, side, and with the same diagnosis group. Repeat injuries were primarily rotator cuff related shoulder pain (44%, 34-55% of repeat injuries), shoulder instability and glenohumeral joint injuries (29%, 20-40%), and other shoulder muscle injuries (23%, 15-33%).

Recurrent injuries were all time-loss injuries, accounting for 25% (17-35%) of all general time-loss injuries. Of these injuries, 70% (48-85%) were shoulder instability and glenohumeral joint injuries and 25% (11-47%) were rotator cuff related shoulder pain. Recurrent injuries required a median of 237 (104-373) days to return to full unrestricted match play, compared to 41 (85-254) days for first time general time-loss injuries (p=0.007).

Exacerbations were noted for 20% (16-24%) of medical attention injuries, requiring additional modification or time off during the recovery process. Exacerbations were primarily noted for rotator cuff related shoulder pain (42% (32-53%) of all injuries with exacerbations; 21% (16-28%) of all rotator cuff related shoulder pain injuries), shoulder instability and glenohumeral joint injuries (37% (27-48%); 29% (21-38%) of shoulder instability and glenohumeral joint injuries), and acromioclavicular joint injuries (6% (3-14%); 18% (8-36%) of acromioclavicular joint injuries).

Fourteen injuries (3% (2-6%) of medical attention injuries, 18% (11-27%) of general time loss injuries) required surgical management, 11 of which followed an exacerbation. 57% (33-79%) were to the dominant side. The average age of players at the time of surgery was 24.3 ± 6.0 years. Injuries requiring surgery were shoulder instability and glenohumeral joint (79% (52-92%) of all injuries requiring surgery; 10% (6-18%) of shoulder instability and glenohumeral joint injuries) and rotator cuff related shoulder pain (21% (8-48%); 2% (1-5%) of rotator cuff related shoulder pain injuries). The median days between initial injury and surgery was approximately 8 months (236 days, IQR 72-485 days) and between surgery and return to full availability was approximately 13 months (388 days, 283-531 days).

Approximately half (54%, 49-59%) of all medical attention injuries resulted in a reduced ability to participate in cricket matches and skills training (the remaining injuries required no modification or affected strength and conditioning training only). Of these injuries requiring modification, the median number of days to return to full unrestricted match play was approximately 2.5 months (73 days, 21-187 days), with 9% (6-14%) of injuries requiring modification persisting for longer than 1 year.

Injuries typically did not result in the player being unable to participate in training or a match, with the exception of sudden onset shoulder instability and glenohumeral joint injuries which resulted in the player being unavailable for a median of 2 weeks (Table 3). When modified activities were required, the modifications continued for between 2 weeks and 6 months. Players typically returned to full unrestricted match play between 1-8.5 months. Gradual onset shoulder instability and glenohumeral joint injuries had a prolonged recovery. Full details of recovery time-frames are presented in Table 3. Within injury categories, injuries to the dominant and non-dominant sides had similar recovery times with the exception of sudden onset rotator cuff related shoulder pain and sudden onset shoulder instability and glenohumeral injuries to the dominant side taking longer to return to full skills training (p=0.022) and unrestricted match play (p=0.040) respectively.

**Table 2.**
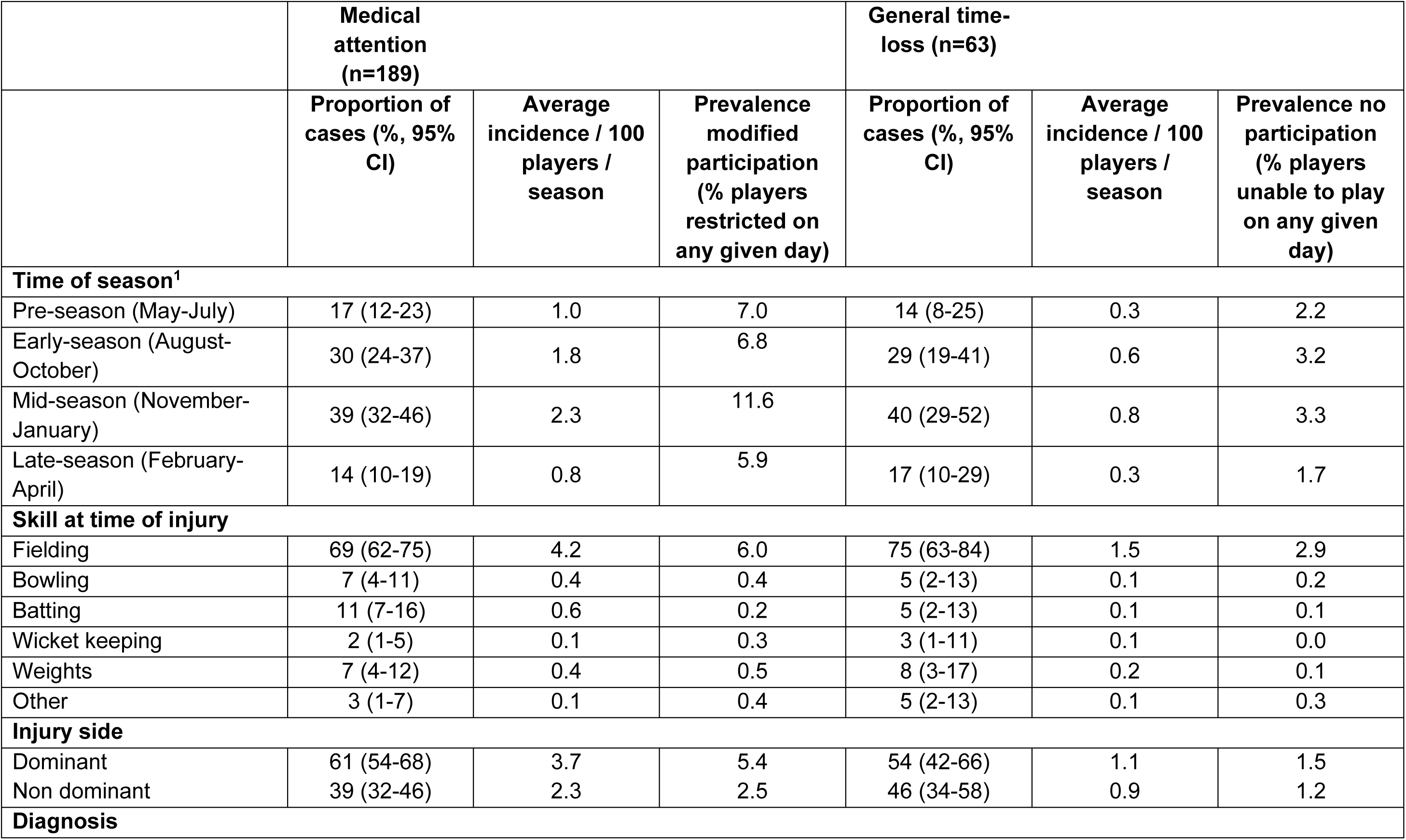
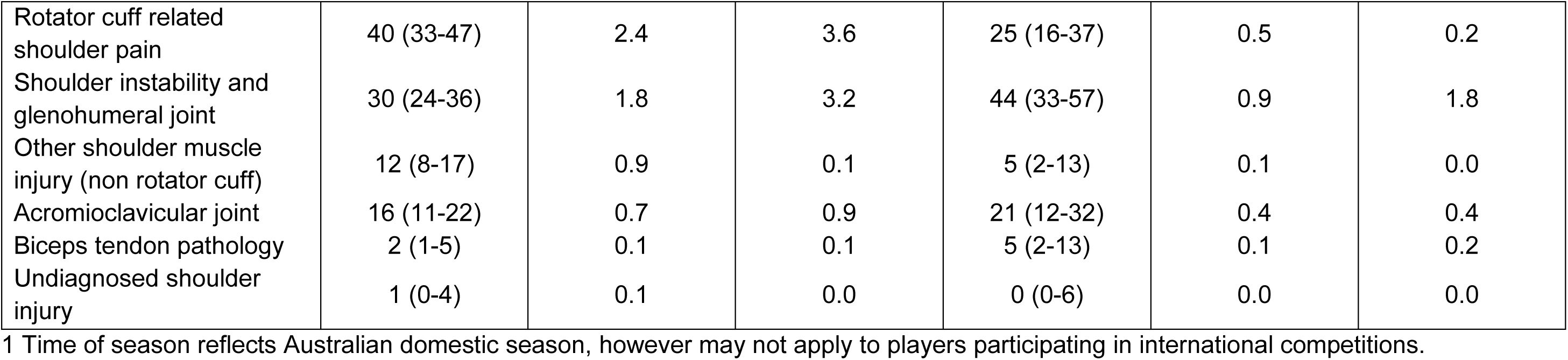
Characteristics of sudden onset shoulder injuries sustained by elite Australian female cricket players over 8 seasons.

**Table 3:**
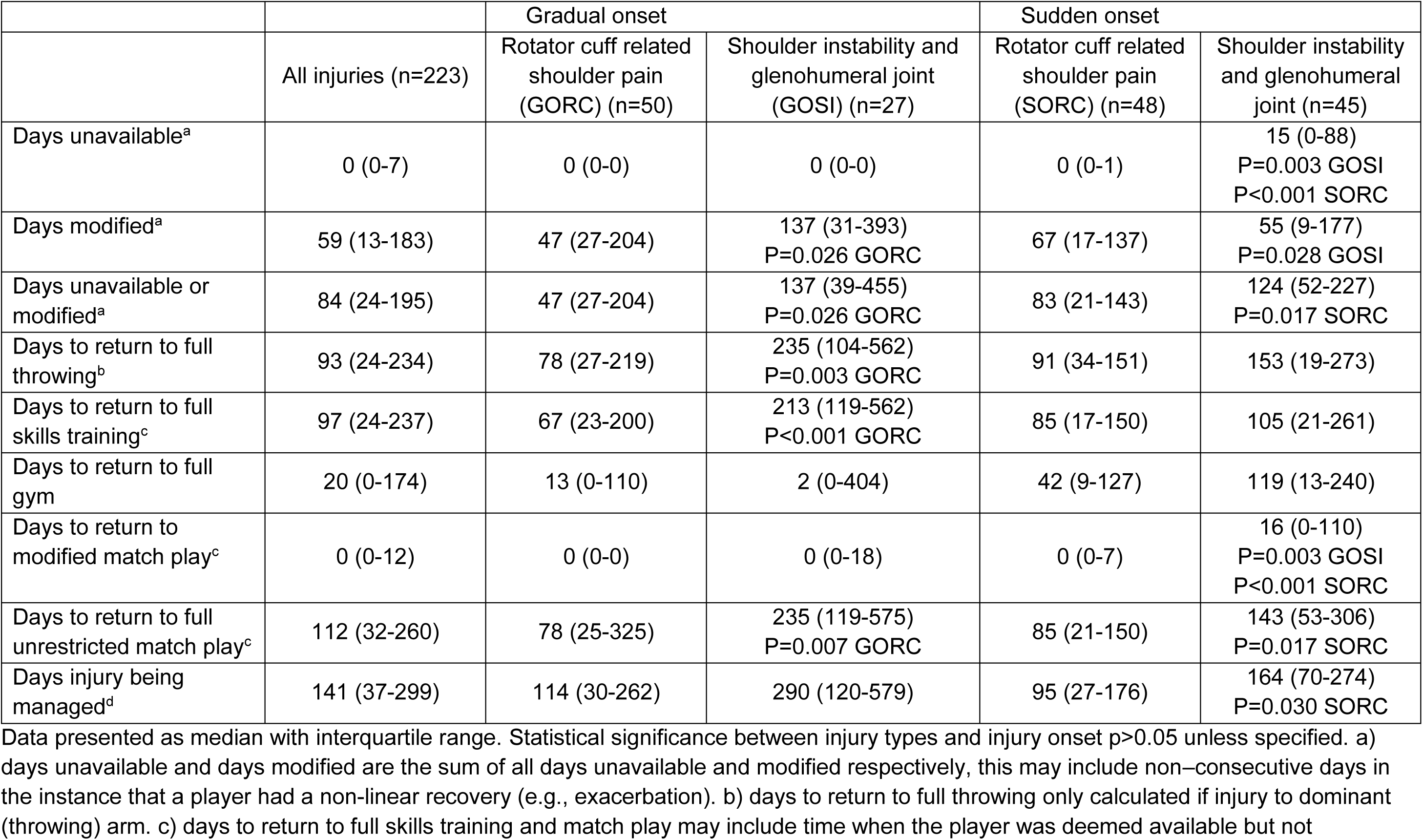
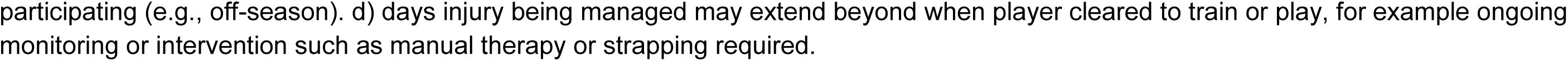
Recovery of predominant shoulder injuries requiring modification and/or time off cricket participation for elite Australian female cricket players over 8 seasons.

|  |  | Gradual onset |  | Sudden onset |  |
| --- | --- | --- | --- | --- | --- |
|  | All injuries (n=223) | Rotator cuff related shoulder pain (GORC) (n=50) | Shoulder instability and glenohumeral joint (GOSI) (n=27) | Rotator cuff related shoulder pain (SORC) (n=48) | Shoulder instability and glenohumeral joint (n=45) |
| Days unavailable<sup>a</sup> | 0 (0-7) | 0 (0-0) | 0 (0-0) | 0 (0-1) | 15 (0-88)<br>P=0.003 GOSI<br>P<0.001 SORC |
| Days modified<sup>a</sup> | 59 (13-183) | 47 (27-204) | 137 (31-393)<br>P=0.026 GORC | 67 (17-137) | 55 (9-177)<br>P=0.028 GOSI |
| Days unavailable or modified<sup>a</sup> | 84 (24-195) | 47 (27-204) | 137 (39-455)<br>P=0.026 GORC | 83 (21-143) | 124 (52-227)<br>P=0.017 SORC |
| Days to return to full throwing<sup>b</sup> | 93 (24-234) | 78 (27-219) | 235 (104-562)<br>P=0.003 GORC | 91 (34-151) | 153 (19-273) |
| Days to return to full skills training<sup>c</sup> | 97 (24-237) | 67 (23-200) | 213 (119-562)<br>P<0.001 GORC | 85 (17-150) | 105 (21-261) |
| Days to return to full gym | 20 (0-174) | 13 (0-110) | 2 (0-404) | 42 (9-127) | 119 (13-240) |
| Days to return to modified match play<sup>c</sup> | 0 (0-12) | 0 (0-0) | 0 (0-18) | 0 (0-7) | 16 (0-110)<br>P=0.003 GOSI<br>P<0.001 SORC |
| Days to return to full unrestricted match play<sup>c</sup> | 112 (32-260) | 78 (25-325) | 235 (119-575)<br>P=0.007 GORC | 85 (21-150) | 143 (53-306)<br>P=0.017 SORC |
| Days injury being managed<sup>d</sup> | 141 (37-299) | 114 (30-262) | 290 (120-579) | 95 (27-176) | 164 (70-274)<br>P=0.030 SORC |

Data presented as median with interquartile range. Statistical significance between injury types and injury onset $p > 0.05$ unless specified. a) days unavailable and days modified are the sum of all days unavailable and modified respectively, this may include non-consecutive days in the instance that a player had a non-linear recovery (e.g., exacerbation). b) days to return to full throwing only calculated if injury to dominant (throwing) arm. c) days to return to full skills training and match play may include time when the player was deemed available but not
participating (e.g., off-season). d) days injury being managed may extend beyond when player cleared to train or play, for example ongoing monitoring or intervention such as manual therapy or strapping required.

Approximately half (52%, 47-56%) of all medical attention shoulder injuries resulted in modifications to cricket-specific skills, and 54% (49-59%) resulted in modifications to any cricket-related activities (including strength and conditioning). The predominant skill requiring modification was fielding (47%, 42-52% of all injuries), followed by bowling (24%, 20-29% of injuries to players with a dominant skill including bowling), and batting (12%, 9-15% of all injuries). 3% (2-5%) of injuries resulted in a player participating in a match in a partial capacity (e.g., an all-rounder that batted but did not bowl).

Cricket-specific fielding modifications included throwing (41%, 36-46% of all injuries), diving (12%, 9-15%), and catching (1%, 1-3%). Throwing modifications included no/avoiding throwing (15%, 12-19%), limiting the volume of throwing (15%, 12-18%), limiting the intensity/distance of throwing (9%, 6-12%), only throwing underarm (6%, 4-8%), and restricting fielding to within the inner circle (4%, 3-7%) (to limit the likelihood of long throws from the outfield). Diving modification included no/avoiding diving (9%, 6-12%), and careful diving (3%, 2-5%). Catching modifications were to avoid catching overhead.

Bowling modifications included no bowling (11%, 8-16% of injuries to players with a dominant skill including bowling), and limiting the number (7%, 5-10%) and intensity (2%, 1-4%) of balls bowled. Batting modifications included avoiding certain types of shots (4%, 2-6% of all injuries) and limiting the volume of batting (1%, 1-3%).

Strength and conditioning modifications included avoiding or modifying aggravating activities (e.g., overhead positions, pushing) (25%, 21-29% of all injuries) and the addition of shoulder rehabilitation exercises (41%, 36-46%). Running activities were avoided or limited for a period of time in 2% (1-3%) of cases. Additional management included specific warm-up exercises (4%, 2-6%), working on technique (2%, 1-3%), and taping (4%, 3-7%). Modifications were similar across injury onset and types of injury. Ongoing monitoring of shoulder strength, range of motion, throwing loads, and pain was indicated in 10% (7-13%) cases once returning to full cricket participation.

## Discussion

This study reviewed 8 seasons of data to understand the incidence and management of shoulder injuries in elite Australian female cricket players. Approximately 13% of players sought medical attention for a shoulder injury each season, with 1 in 5 requiring time off cricket. At any given time, 2-3% of players were unable to fully participate and hence perform as a result of a shoulder injury. This study confirms previous studies that shoulder injuries pose a considerable burden for elite female cricket players^17^,and offers additional insight into the types of injuries and which players are more at risk.

Gradual onset injuries contributed just over half of all medical attention injuries, with the dominant arm of pace bowlers most affected. Intuitively, the dominant arm experiences higher volumes and is a consistent risk factor identified amongst female athletes participating in overhead sports.^18^ In addition to the load of the pace bowling action on the shoulder, pace bowlers tend to field in the outfield when not bowling which adds a high throwing load to the shoulder.^14^ Both the pace bowling action and throwing action cause a large distraction force at the shoulder joint which the rotator cuff must resist.^12,19,20^. Tensile failure of the rotator cuff may result in rotator cuff pathology.^19^ High chronic loads^12,21–23^ and/or a high acute load with few rest days^23^ may lead to a gradual onset shoulder injuries. Therefore, bowling and throwing load regulation may be implemented to reduce the risk of shoulder injury, supported by increased strength and sufficient mobility.^24^

Throwing requires both stability and mobility of the shoulder. To achieve this, overhead throwing athletes have been observed to develop increased humeral retroversion as an adaptation to the demands of throwing.^25^ However, this adaptation was not observed in a study of 105 elite male cricket players, leading the authors to conclude that cricketers do not present with the classical ‘throwing shoulder’.^26^ The authors attributed this lack of adaptation to less throwing in junior cricketers and overall weakness compared to other throwing athletes.^26^ A similar lack of adaptation and strength for throwing may explain the specific shoulder injuries seen in this elite female cohort. Females may also have increased joint laxity and an increased susceptibility to tendinopathy due to sex-specific inflammatory mediators and hormones,^27^ hence throwing load regulation, strength, and mobility interventions may need to be sex-specific.

Greater capsular laxity of females compared to males^27^ may also contribute to an increased susceptibility to traumatic injuries. Sudden onset injuries in this study were often the result of diving to field the ball. Impact may have been directly to the shoulder or indirectly through force transmitted through the arm.^28^ Diving to field a ball is an important skill in cricket, hence a feasible risk reduction strategy is teaching players diving/falling techniques to lessen the impact on the shoulder and other vulnerable structures.^28,29^ Given just over half of all sudden onset injuries occurred outside of matches, there is opportunity to implement additional strategies in training and warm-up to further reduce the risk of shoulder injury.

The second purpose of this study was to describe the management of shoulder injuries in elite female cricket players, including modifications and time to return to specific activities. 20% (16-24%) of shoulder injuries required any time off cricket, primarily sudden onset shoulder instability and glenohumeral joint injuries. In most cases, the shoulder injury was managed with modifications whilst continuing to participate in cricket training and matches. A key modification was to limit throwing, which was consistent with what has been previously described in elite male cricket players.^9^ In elite female cricket, players operating at reduced capacity are still valuable members of the team if they can perform their dominant skill (batting and/or bowling). However, other team members must accommodate the injured player’s modifications which in-turn may place other players at increased risk of shoulder injuries (e.g., an injured player restricted to field in the inner field requires other players to field and throw from the outfield).

Approximately one in five cases experienced one or more exacerbations during the recovery process. In these cases, continuing to play, albeit with modifications to protect the injured shoulder, may not have provided the optimum opportunity for rehabilitation.

The burden of injuries extends beyond the performance implications of a player unable to fully participate in cricket. The management details also highlighted the addition of rehabilitation exercises, warm-up exercises, taping, and ongoing monitoring which place additional burden on injured players and support staff throughout the extended recovery period.

Long-term implications of shoulder injury are another potential source of injury burden. Previous shoulder injury is associated with subsequent shoulder problems including pain and reduced function and performance^30–32^ and may also be a risk factor for subsequent injury.^33^ The impact of previous injury may have contributed to the 9% of the player cohort who experienced more than one medical attention shoulder injury during the 8 year study period. Approximately one in four general time-loss injuries were a recurrence of a previous injury, with prolonged recovery and in some cases surgery an additional implication.

A limitation of this study is that for an injury to be included, the player had to have sought medical attention. It is possible that lower-level modifications may have been mentioned in consultation with the player but not recorded, or the player may have chosen to self-modify some activities without reporting this to staff or having it recorded. Research using self-report can increase the capture of traditional injury reporting by medical staff.^33^ For example, research using player self-reported pain have found high rates of players affected by shoulder pain despite continuing to participate in their sport: 60% of female softball pitchers;^31^ almost half of non-injured college athletes,^32^ and; 38% of a small sample of female local-international-level water polo players.^34^ An additional limitation is that shoulder injuries were only included in this study if they were the result of cricket participation. This excluded 18 injuries which occurred independent of cricket participation, however required management by medical staff within cricket and impacted cricket participation. In light of these limitations, the overall burden of shoulder injuries in elite female players may be higher than reported in this study. Future research may take a mixed-methods approach to expand the capture of injuries.

## Conclusion

This study demonstrated that shoulder injuries are common and a considerable burden in elite female cricket players. Despite only one in five injuries resulting in a player being unavailable to play or train, there is considerable burden on the player, their team, and support staff to accommodate modifications. Risk reduction strategies may include: throwing load regulation; bowling load regulation particularly in pace bowlers; implementing exercise programs to target shoulder strength, power, and mobility, and; coaching throwing, diving, and falling technique. Efforts should also be directed at enhancing rehabilitation to reduce the risk of exacerbation and recurrent injury.

### Practical implications

- Approximately thirteen percent of elite female cricket players are unable to fully participate in training and/or match play due to a shoulder injury each season.
- Players typically continue to train and play with modifications to accommodate their shoulder injury over several months, potentially impacting individual and team performance.
- Practitioners should consider strategies to reduce the burden of shoulder injuries, such as load regulation, exercise programs, technique coaching, and enhancing rehabilitation.

## Data Availability

Non-identifiable data are available upon reasonable request

## Acknowledgements

This research was supported in-kind by Cricket Australia. The authors thank the sport science and sport medicine staff who assessed players and recorded data throughout the study period to enable this retrospective study.

## Data sharing

Non-identifiable data are available upon reasonable request.

## Ethical approval

Ethics approval was granted from La Trobe University Human Ethics Committee (HEC20058) which waived the need for individual informed consent.

## Author contributions

GP and PD contributed to conceptualization, data curation, investigation, validation, writing-review and editing. AS contributed to conceptualisation, data curation, formal analysis, investigation, methodology, project administration, visualisation, writing-original draft, writing-review and editing. methods, data collection and curation, analysis, and drafting of the manuscript. KB contributed to conceptualization, validation, supervision, writing-review and editing. KS contributed to validation, supervision, writing-review and editing.

## References

1. Lin DJ, Wong TT, Kazam JK. Shoulder injuries in the overhead-throwing athlete: epidemiology, mechanisms of injury, and imaging findings. Radiol. 2018;286(2):370–387. doi:10.1148/radiol.2017170481

2. Orchard JW, Ranson C, Olivier B, et al. International consensus statement on injury surveillance in cricket: a 2016 update. Br J Sports Med. Oct 2016;50(20):1245–1251. doi:10.1136/bjsports-2016-096125

3. Goggins L, Peirce N, Ranson C, et al. Injuries in England and Wales elite men’s domestic cricket: A nine season review from 2010 to 2018. J Sci Med Sport. 2020;23(9):836–840. doi:10.1016/j.jsams.2020.03.009

4. Orchard JW, Inge P, Sims K, et al. Comparison of injury profiles between elite Australian male and female cricket players. J Sci Med Sport. 2023/01/01/ 2023;26(1):19–24. doi:10.1016/j.jsams.2022.12.002

5. Goggins L, Warren A, Smart D, et al. Injury and player availability in women’s international pathway cricket from 2015 to 2019. Int J Sports Med. Oct 2020;41(13):944–950. doi:10.1055/a-1192-5670

6. Frost WL, Chalmers DJ. Injury in elite New Zealand cricketers 2002–2008: descriptive epidemiology. Br J Sports Med. 2014;48(12):1002–1007. doi:10.1136/bjsports-2012-091337

7. Perera NKP, Kountouris A, Kemp JL, Joseph C, Finch CF. The incidence, prevalence, nature, severity and mechanisms of injury in elite female cricketers: A prospective cohort study. J Sci Med Sport. 2019;22(9):1014–1020. doi:10.1016/j.jsams.2019.05.013

8. Warren A, Dale S, McCaig S, Ranson C. Injury profiles in elite women’s T20 cricket. J Sci Med Sport. 2019;22(7):775–779.

9. Ranson C, Gregory PL. Shoulder injury in professional cricketers. Physical Ther Sport. 2008/02/01/ 2008;9(1):34–39. doi:10.1016/j.ptsp.2007.08.001

10. Arora M, Shetty SH, Dhillon MS. The shoulder in cricket: What’s causing all the painful shoulders? J Arthroscopy Joint Surg. 2015/05/01/ 2015;2(2):57–61. doi:10.1016/j.jajs.2015.06.003

11. Walter S, Petersen C, Basu A. Quantifying injuries among New Zealand cricket fast bowlers: A 12-month retrospective injury surveillance. NZ J Sports Med. 2021;48(2):46.

12. Murphy MC, Chivers P, Mahony K, Mosler AB. Risk factors for dominant-shoulder injury in elite female Australian cricket players: A prospective study. Transl Sports Med. 2020;3(5):404–414. doi:10.1002/tsm2.158

13. Goggins L, Williams S, Griffin S, Langley B, Newman D, Peirce N. English and Welsh men’s domestic cricket injury risk by activity and cricket type: A retrospective cohort study from 2010 to 2019. J Sci Med Sport. 2024;27(1):25–29. doi:10.1016/j.jsams.2023.07.013

14. Bell-Jenje T. Incidence, nature and risk factors in shoulder injuries of national academy cricket players over 5 years-a retrospective study. South African J Sports Med. 2005;17(4):1–7.

15. Walter S, Petersen C, Basu A. Are there any differences in shoulder muscle strength and range of motion between fast bowlers with and without shoulder pain? NZ J Sports Med. 2021;48(1)

16. Rae K, Orchard J. The Orchard Sports Injury Classification System (OSICS) Version 10. Clin J Sport Med. 2007;17(3)doi:10.1097/JSM.0b013e318059b536

17. Jacobs J, Olivier B, Dawood M, NK PP. Prevalence and incidence of injuries among female cricket players: a systematic review and meta-analyses. JBI Evidence Synthesis. 24/12/2021 2021;doi:10.11124/jbies-21-00120

18. Steele MC, Lavorgna TR, Ierulli VK, Mulcahey MK. Risk Factors for shoulder injuries in female athletes playing overhead sports: A systematic review. Sports Health. Jun 20 2024:19417381241259987. doi:10.1177/19417381241259987

19. Werner SL, Gill TJ, Murray TA, Cook TD, Hawkins RJ. Relationships between throwing mechanics and shoulder distraction in professional baseball pitchers. Am J Sports Med. 2001;29(3):354–358. doi:10.1177/03635465010290031701

20. Stuelcken MC, Ginn KA, Sinclair PJ. Shoulder strength and range of motion in elite female cricket fast bowlers with and without a history of shoulder pain. J Sci Med Sport. Nov 2008;11(6):575–80. doi:10.1016/j.jsams.2007.06.007

21. Asker M, Brooke HL, Waldén M, et al. Risk factors for, and prevention of, shoulder injuries in overhead sports: a systematic review with best-evidence synthesis. Br J Sports Med. 2018;52(20):1312–1319. doi:10.1136/bjsports-2017-098254

22. Mehta S, Tang S, Rajapakse C, Juzwak S, Dowling B. Chronic workload, subjective arm health, and throwing injury in high school baseball players: 3-year retrospective pilot study. Sports Health. 2022;14(1):119–126. doi:10.1177/19417381211055142

23. Saw R, Dennis RJ, Bentley D, Farhart P. Throwing workload and injury risk in elite cricketers. Br J Sports Med. 2011;45(10):805–808. doi:10.1136/bjsm.2009.061309

24. Paul J, Brown SM, Mulcahey MK. Injury prevention programs for throwing injuries in softball players: a systematic review. Sports Health. 2021;13(4):390–395. doi:10.1177/1941738120978161

25. Downar JM, Sauers EL. Clinical measures of shoulder mobility in the professional baseball player. Journal of athletic training. 2005;40(1):23–29.

26. Dutton M, Tam N, Brown JC, Gray J. The cricketer’s shoulder: Not a classic throwing shoulder. Physical Ther Sport. 2019;37:120–127. doi:10.1016/j.ptsp.2019.03.014

27. Wessel LE, Eliasberg CD, Bowen E, Sutton KM. Shoulder and elbow pathology in the female athlete: sex-specific considerations. J Shoulder Elbow Surg. 2021;30(5):977–985. doi:10.1016/j.jse.2020.10.020

28. Krogsgaard MR, Safran MR, Rheinlaender P, Cheung E. Preventing shoulder injuries. In: Bahr R, Engebretsen L, eds. Handbook of sports medicine and science: Sports injury prevention. John Wiley & Sons; 2009:134–152:chap 9.

29. Steffen K, Andersen TE, Krosshaug T, et al. ECSS Position Statement 2009: Prevention of acute sports injuries. Eur J Sport Sci. 2010;10(4):223–236.

30. Alonso-Muñoz MB, Calvache-Mateo A, Martín-Núñez J, et al. Musculoskeletal, functional and performance impairment in female overhead athletes with a previous shoulder injury. Healthcare. 2023;12(1):21.

31. Sauers EL, Dykstra DL, Bay RC, Bliven KH, Snyder AR. Upper extremity injury history, current pain rating, and health-related quality of life in female softball pitchers. J Sport Rehab. 2011;20(1):100–114. doi:10.1123/jsr.20.1.100

32. Soldatis JJ, Moseley JB, Etminan M. Shoulder symptoms in healthy athletes: a comparison of outcome scoring systems. J Shoulder Elbow Surg. 1997;6(3):265–271. doi:10.1016/S1058-2746(97)90015-X

33. Whalan M, Lovell R, Sampson JA. Do Niggles Matter? - Increased injury risk following physical complaints in football (soccer). Sci Med Football. 2020/07/02 2020;4(3):216–224. doi:10.1080/24733938.2019.1705996

34. Girdwood M, Webster M. Quantifying the Burden of Shoulder and Hip Pain In Water Polo Players Across Different Playing Levels. Int J Sports Physical Ther. 2021;16(1):57–63. doi:10.26603/001c.18801

